# Interventions that help to improve spontaneous adverse drug reaction reporting by patients: a protocol of a scoping review

**DOI:** 10.1101/2024.02.27.24303435

**Authors:** Romina P Martinelli, M Gabriela Papazian

## Abstract

When a new medical product is released to the market, a continuous pharmacovigilance process is initiated to guarantee patient safety by collecting and analyzing adverse drug reaction (ADR) reports. These ADRs are noxious and unintended responses to a medicine and are collected and analyzed in databases like EudraVigilance to determine safety profiles of the products and signal detection. The spontaneous reporting of suspected ADR could be performed by both health care workers and patients/consumers. However, the under-reporting is well known. Different initiatives have been developed to encourage reporting by health professionals, however, further work is required to support patients in taking a more active role in reporting adverse drug reactions. In this context, we will conduct a Scoping Review of the literature to inquire about what is known about actions or strategies to improve pharmacovigilance engagement by patients. We will conduct searches in MEDLINE/PubMed, Scielo, Latindex, DOAJ, CINAHL, LILACS, IAM, IMEMR, IMSEAR, WPRO, and Cochrane Library databases. Two reviewers will screen all identified records for relevance. Conflicts between reviewers will be solved by consensus. We will chart the data using data-charting forms. Findings will be reported according to PRISMA for Scoping Reviews (PRISMA-ScR). No quality assessment will be performed.

## 1 Rationale

Immediately after a new medical product is released to the market, a constant pharmacovigilance process is set in motion. This exhaustive system has the purpose of ensuring patient safety by collecting and analyzing adverse drug reaction (ADR) reports.

These ADRs, defined as “noxious and unintended responses to a medicine”^1^, should be reported to EudraVigilance database designed to collect and analyze the data^2^. This valuable information contributes with drugs safety profiles and signal detection. Considering a signal in pharmacology a “Reported information on a possible causal relationship between an adverse event and a drug, the relationship being previously unknown or incompletely documented^3^.

The spontaneous reporting of suspected ADR could be performed by both health care workers and patients/consumers. However, the under-reporting is well known. Hazell (2006)^4^ showed median under-reporting rate of 95% in a systematic review published in 2006. Consequently, under-reporting could delay signal detection, compromising drug safety.

In Europe, during 2022, experienced a reduction of the 17% in the number reports submitted directly by patients/consumers compared to the previous year^5^. In 2021, there had been a huge increase in reports due to the launch of covid-19 vaccines. While the number is still high compared to 2020, this downward trend should draw our attention (Figure 1).

**Figure 1.**
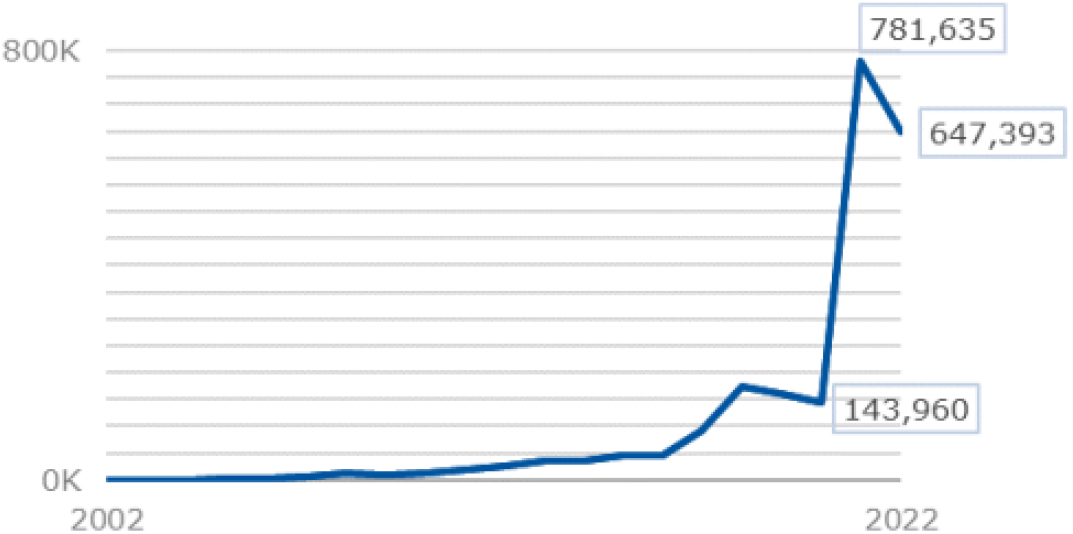
Trend of ADR reports from patient and consumers reported to EudraVigilance. Reproduced from *European Medicines Agency (EMA). 2022 Annual Report on EudraVigilance for the European Parliament, the Council and the Commission EMA/900566/2022*.

During the last few years, different initiatives have been developed to encourage reporting by health professionals^6^. However, further work would be required to support patients in taking a more active role in reporting adverse drug reactions^7^. For this reason, the present scoping review aims to explore the existing literature to provide an overview of actions or strategies to improve pharmacovigilance engagement in patients.

## 2 Objectives

In this review, we aim to inquire about what is known about actions or strategies to improve pharmacovigilance engagement in patients to promote ADR reporting.

The scoping review will be carried out to answer the following question: What is known in the existing literature about the interventions proposed and/or carried out on patients to encourage ADR reporting?

The main objectives of this review are:

i. To screen the published literature reporting interventions on patients/consumers to improve ADR reporting.
ii. To map the type of patient/consumers involved.
iii. To summarize the effectiveness of the interventions carried out.

## 3 Methods

To develop the scoping review protocol, the methodological framework introduced by Arksey and O’Malley^8^ was followed, and it was reported according to the PRISMA for Scoping Reviews (PRISMA-ScR) ^9^.

### Stage 1: identifying the research question

To formulate the research question, the “PICO” methodology was used.

#### Population

Adult patients / Healthcare consumer

#### Intervention

Activities/ interventions/ actions to improve patient engagement or ADR reporting.

#### Comparison

NA

#### Outcomes

NA

### Stage 2: identifying relevant studies

The search will be manually conducted in MEDLINE/PubMed, Scielo, Latindex, DOAJ, CINAHL, LILACS, IAM, IMEMR, IMSEAR, WPRO, and Cochrane Library databases. The search strategy will include Medical Subject Headings (MeSH) terms, and their relevant synonyms joined with Boolean operators aligned with each Database. Searches will not be restricted by publication date, place, or type of study. Systematic reviews and meta-analyses will be considered as additional sources of primary studies. There will be no language restrictions.

The general search strategy proposed, will include the following terms, and will be adapted for each search engine:

> (adverse drug reaction* report* OR adr report* OR adverse drug event* OR report* OR side effect* OR report* pharmacovigilance) AND (improv* OR motivat* OR incentiv* OR increas* OR interven* OR educat* OR train* OR action* OR action* OR program* OR feedback* OR system* OR trend*) AND (patient* OR consumer* OR population* OR citizen*)

Search results will be downloaded in Comma-Separated Values (CSV) format on a single day.

### Stage 3: study selection

After removing duplicates, two researchers (MGP and RPM) will screen the results at the title/ abstract level and second at the full-text level independently. Discrepancies will be resolved by consensus between the two reviewers. There will be no language restrictions.

#### Inclusion criteria

Publications reporting on actions/programs to improve patient engagement or ADR reporting will be included. There is no restriction regarding study design.

#### Exclusion criteria

Publications reporting on actions/programs to improve ADR reporting by health professionals will be excluded.

#### Stage 4: charting the data

Data extraction will be independently performed by two researchers (MGP and RPM), and discrepancies will be resolved by consensus.

We developed a data charting form with the following categories: Citation, Country, Study Design, Target Population, Type of Intervention, Aim, Follow Up Duration, Study Outcome, PV action (if apply), Founding. This form could be correct after being tested on 10 random studies.

### Stage 5: collating, summarizing, and reporting the results

The PRISMA flowchart will be used to present the study selection process^10^. Data will be presented in descriptive form.

## 4 Ethics and dissemination

This review does not require ethical approval. The findings will be submitted for publication in an international meeting and scientific journal.

## Data Availability

All data produced in the present work are contained in the manuscript

## 5 Contributions of authors

MGP and RPM drafted the protocol. MGP and RPM developed the search strategy and conducted electronic searches. PCP helped to develop the protocol.

